# Applications of Artificial Intelligence in Neurosurgical Education: A Scoping Review

**DOI:** 10.1101/2025.02.07.25321681

**Authors:** Hector Julio Piñera-Castro, Christian Borges-García

## Abstract

**Background:** Artificial intelligence (AI) has transformed medical education through optimized instruction, competency assessment, and personalized learning. Its integration into neurosurgical education, given the field’s complexity and precision demands, warrants comprehensive exploration.

**Objective:** To systematically evaluate AI applications in neurosurgical education.

**Methods:** A scoping review adhering to PRISMA-ScR guidelines was conducted. A Scopus search (up to May 2024) identified 23 eligible studies. Inclusion criteria encompassed peer-reviewed observational or experimental studies on AI in neurosurgical education. Narrative synthesis categorized findings into key domains.

**Results:** Four main key areas emerged: performance in board examinations and ethical considerations, simulation-based training and tutoring, performance/skills/expertise analysis and assessment, and other applications. In board examinations, GPT-4 outperformed prior models and junior neurosurgeons in text-based questions but lagged in image-based tasks. Simulation training utilized neural networks to classify expertise and deliver individualized feedback, though rigid metrics risked oversimplifying skill progression. Machine learning models assessed surgical performance, identifying metrics. Other innovations included AI-generated academic content, neuroanatomical segmentation, and instrument pattern analysis. Ethical concerns highlighted risks of overreliance, image-processing limitations, and the irreplaceable role of clinical intuition. Technical challenges included dataset biases and simulation realism.

**Conclusions:** AI enhances neurosurgical education through knowledge assessment, simulation feedback, and skill evaluation. However, integration requires addressing ethical dilemmas, improving multimodal data processing, and ensuring human-AI collaboration. Continuous model refinement, expanded datasets, and hybrid curricula combining AI analytics with expert mentorship are critical for safe, effective implementation. This evolution promises to elevate training quality while preserving the indispensable value of hands-on experience in neurosurgical practice.

## INTRODUCTION

Artificial intelligence (AI) is an interdisciplinary domain dedicated to enabling computers to execute tasks traditionally associated with human cognition, such as perception, reasoning, problem-solving, and planning. Central to AI are machine learning (ML) and data analytics, which empower machines to emulate human cognitive processes and address complex, ambiguous problems with intentionality, intelligence, and adaptability. AI finds applications across various sectors, notably within the medical field.^(1–3)^

AI has a significant impact on medical education by transforming educational paradigms and enhancing learning experiences. AI technology ensures the safety and consistency of medical experiments, fosters diagnostic reasoning, and heightens humanistic care awareness among students. It amalgamates resources, aids instruction, simulates clinical environments, monitors teaching quality, facilitates self-evaluation, and enables personalized learning. Additionally, AI equips healthcare professionals with superior skills and knowledge, leading to improved practical competencies and better patient outcomes. The provision of immediate and interactive feedback reinforces knowledge, and overall, AI elevates the quality of medical education, inspiring innovative applications within the field.^(4–8)^

Given the interdisciplinary nature and transformative potential of AI, its integration into neurosurgical education merits thorough examination. The complexity and precision required in neurosurgical training amplify the potential benefits of AI, which includes enhancing educational paradigms, ensuring the safety and consistency of training methods, and providing adaptive learning experiences.

Therefore, it is pertinent to ask: What are the current applications of AI in neurosurgical education? This review systematically evaluates these applications, consolidating existing knowledge and highlighting best practices.

By doing so, it seeks to inspire innovative applications and inform future research, ultimately aiming to heighten the proficiency and preparedness of neurosurgeons through AI-enhanced education.

The relevance of these findings extends to neurosurgical students and residents, medical educators and institutions, policymakers, and AI developers and researchers. AI offers tailored feedback, skill development, and adaptive learning experiences for students and residents. For educators and institutions, AI provides supplementary evaluative tools and personalized training plans. Policymakers must address ethical concerns to ensure responsible use, while developers and researchers should refine AI models, enhance datasets, and collaborate with medical experts to further AI’s applications in neurosurgical education.

## METHODS

This scoping review adhered to the Preferred Reporting Items for Systematic Reviews and Meta-Analyses (PRISMA),^(9)^ complemented with the PRISMA for Scoping Reviews (PRISMA-ScR)^(10)^ and the PRISMA for Abstracts (PRISMA-A) extensions.^(11)^

### Eligibility criteria

The inclusion criteria comprised observational or experimental original studies that investigated the application of AI in the context of neurosurgical education. These studies were required to be published in peer-reviewed journals and could encompass various methodological approaches, including but not limited to randomized controlled trials, cohort studies, case-control studies, and cross-sectional studies.

Only original observational or experimental were included in order to prioritize empirical, data-driven evidence on AI’s educational applications. Excluding reviews, editorials, or theoretical works ensured a focus on primary research with measurable outcomes, enhancing rigor and relevance. Peer review guaranteed methodological validity.

The exclusion criterion was limited to studies not written in English or Spanish. English is the dominant language of peer-reviewed scientific literature, ensuring access to the majority of high-quality, globally relevant research in AI and neurosurgery. Including Spanish accommodates contributions from regions with active neurosurgical research communities (e.g., Latin America, Spain) without overextending translation resources. This approach balances inclusivity with logistical constraints (e.g., time, cost, availability of translators for less common languages), while minimizing the risk of omitting critical evidence, as most impactful studies in this field are published in these languages.

### Information Sources and Search Strategy

We conducted a literature search in Scopus, last accessed on May 25^th^, 2024. No date or language restrictions were applied initially. In addition, we reviewed reference lists of relevant articles to identify any further eligible studies.

Scopus was selected as the sole database for this scoping review due to its multidisciplinary scope, ensuring comprehensive coverage of peer-reviewed journals. Its rigorous quality control minimizes inclusion of non-peer-reviewed sources. Though omitting specialized databases like PubMed risks excluding niche studies, Scopus’s breadth across biomedical fields sufficiently captures the review’s focus while streamlining the process, making it ideal for exploring the landscape of AI applications in neurosurgical education without compromising rigor.

### Search Strategy

The full search strategy involved using a combination of terms related to “neurosurgery” and “artificial intelligence”, within the article title, abstract, and keywords: *(Neurosurg* OR “Neurologic* Surgery”) AND (“Artificial Intelligence” OR “AI” OR “Computer Reasoning” OR “Machine Intelligence” OR “Computational Intelligence” OR “Computer Vision System*” OR “Computer Knowledge Acquisition” OR “Computer Knowledge Representation*”)*. We decided not to include terms related to education due to the extensive range of possibilities. Instead, we preferred to select the articles pertinent to the topic manually. The filter applied limited the results to original research articles.

### Selection Process

Two independent reviewers screened the titles and abstracts to identify studies that might be eligible. The full texts of studies deemed potentially relevant were retrieved and independently assessed for eligibility by the same reviewers. Any discrepancies in judgment were resolved through discussion or by consulting a third reviewer.

### Data Collection Process

Data extraction was conducted independently by two reviewers using a standardized form. Any discrepancies were resolved through consensus. Attempts were made to contact the authors of the studies for clarifications or additional data as necessary. No automation tools were employed in the data collection process.

### Data items

The following data were extracted: publication year, author(s), study design, participants, objective, AI model(s)/algorithm(s)/technique(s) utilized, main results, contributions, limitations, and conclusions.

### Risk of Bias Assessment and Effect Measures

Due to the heterogeneity of the study designs, interventions, and outcomes, effect measures were not utilized and a risk of bias assessment was not performed. The substantial variability in study designs, AI applications, and reported outcomes made it challenging to aggregate the data quantitatively. Additionally, the primary aim of this review was to provide a broad overview of the current landscape and innovative uses of AI in neurosurgical education rather than to quantify the effectiveness of specific interventions.

### Data Synthesis

A narrative synthesis approach was employed to collate and summarize the findings across the included studies. This approach allowed for a comprehensive exploration of the various applications of AI in neurosurgical education, highlighting common themes, innovations, and challenges reported in the literature. The narrative synthesis provided a rich contextual understanding of the impact and potential of AI in this field without relying on quantitative effect measures.

## RESULTS AND DISCUSSION

The application of the search strategy yielded 362 reports. Following a screening of titles and abstracts, 303 studies were identified as potentially eligible, of which only 275 were successfully retrieved. After conducting an eligibility assessment, 252 reports were excluded for various reasons, resulting in 23 studies being included in the review. The search and study selection process followed the procedure outlined in **Figure 1**.

**Figure 1.**
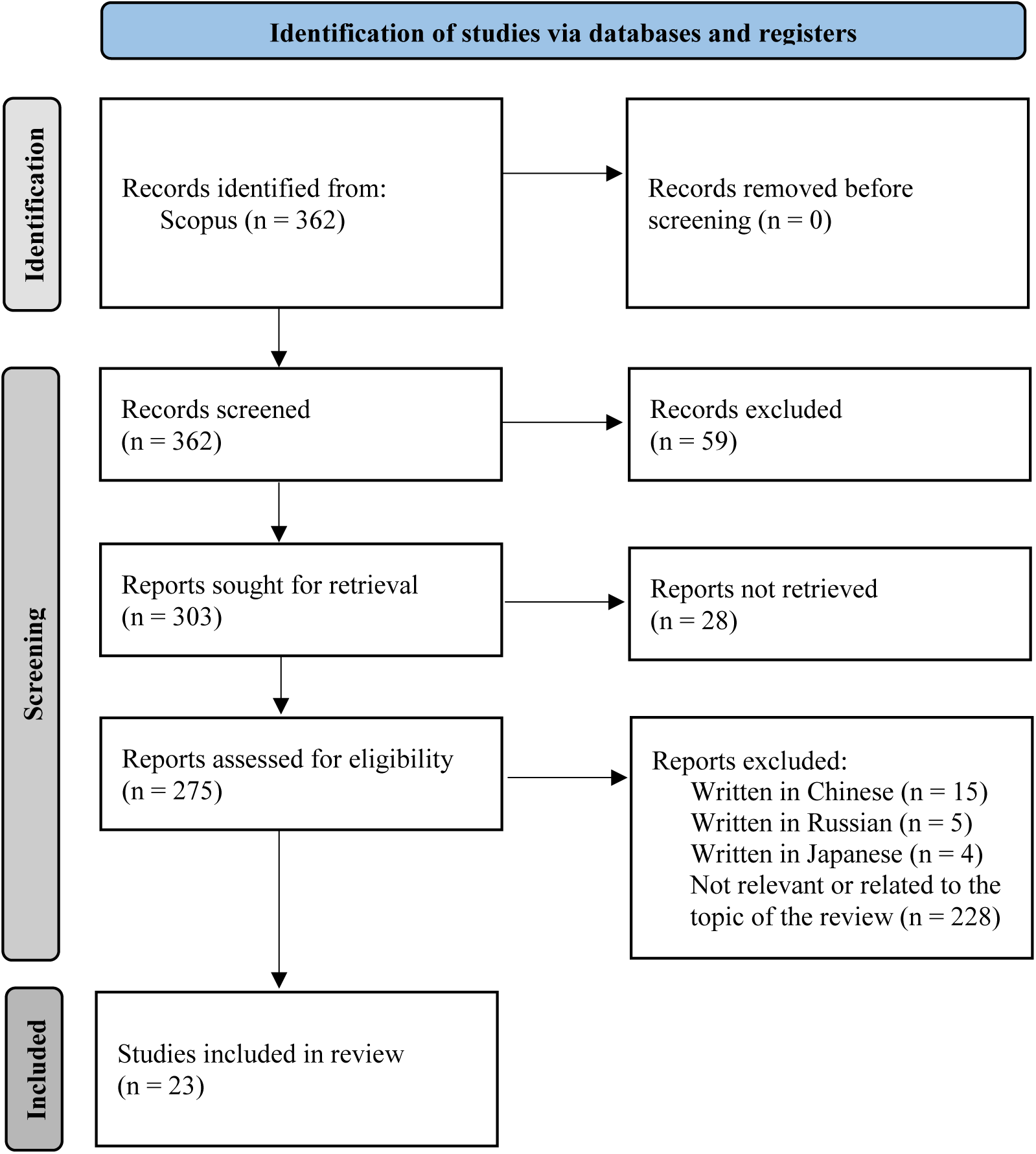
Flowchart of the Search and Study Selection Processes.

### General overview

This review covers four key areas based on the synthesis of evidence from the selected articles: performance in neurosurgical board examinations and ethical considerations, neurosurgical simulation-based training and tutoring, neurosurgical performance/skills/expertise analysis and assessment, and other applications in neurosurgical education (**Table 1**).

**Table 1.** Applications and Limitations of AI in Neurosurgical Education.

| Key areas | Application(s) | Limitations |
| --- | --- | --- |
| Performance in neurosurgical board examinations and ethical considerations<sup>(12–19)</sup> | Answer neurosurgical board examination questions, particularly text-based questions. | • Inferior performance in practical exams and complex clinical judgment scenarios.<br>• Disparities in accuracy, especially in licensing exams.<br>• Challenges in detecting and processing information from images.<br>• Difficulty in addressing open-ended questions and real-world neurosurgical complexity.<br>• Risk of overreliance on AI, leading to demotivation and loss of fundamental clinical skills. |
| Neurosurgical simulation-based training and tutoring<sup>(20–24)</sup> | • Analyze performance metrics during surgical simulations.<br>• Learn the set of metrics for a specific procedure that comprise surgical expertise.<br>• Classify participants by experience level using neural networks and other algorithms.<br>• Provide individualized and quantitative feedback based on metrics obtained in simulations.<br>• Offer real-time audiovisual and metric evaluations.<br>• Assess the impact of various feedback modalities (numeric, visual, visuospatial) on performance improvement.<br>• Develop personalized training plans.<br>• Identify learning patterns. | • Binary classifications are too general and fail to capture nuances in technical competencies.<br>• Difficulties in comprehensively evaluating interdisciplinary skills and team collaboration aspects.<br>• Biases in evaluated metrics, as high values in certain parameters can mask deficiencies in other critical aspects.<br>• Potential negative effects on participants' skills, as AI-enhanced training curriculum were limited to post-exercise comments, potentially lacking important contextual components. |
| Neurosurgical performance/skills/expertise analysis and assessment<sup>(25–31)</sup> | • Evaluate competencies and surgical skills during simulated procedures.<br>• Use classification algorithms to categorize participant performance.<br>• Analyze specific metrics (e.g., contact time, contact length, inter-instrument distance) to assess technical proficiency. | • Small datasets hinder model generalization to broader contexts or new scenarios.<br>• Simulation scenarios fail to reproduce accurately the biomechanical properties of real tissues, affecting evaluative precision. |
|  | Identify patterns across different expertise levels.<br>Identify the surgical task being performed in real-time. | Difficulty in holistically integrating all necessary competencies in neurosurgical practice, especially complex behavioral aspects. |
| Other applications in neurosurgical education<sup>(32–34)</sup> | Generate clinical cases and multiple-choice questions for academic purposes.<br>Draft academic articles.<br>Implement neuroanatomical segmentation algorithms based on convolutional neural networks for image analysis and structure identification.<br>Analyze surgical instrument usage patterns using computer vision techniques.<br>Use measures like Shannon entropy to predict surgical procedure success. | Generate academic content errors, such as multiple-choice questions with multiple correct answers or inaccurate content.<br>Limit the accuracy of neuroanatomical segmentation algorithms due to the imprecision of manual reference labels.<br>Face challenges in analyzing surgical instrument usage patterns, especially in heterogeneous procedures where entropy may vary without reflecting actual success or failure. |

Source: Elaborated by the authors.

### Performance in Neurosurgical Board Examinations and Ethical Considerations

The review encompassed eight studies^(12–19)^ that evaluated the performance of various AI chatbots in neurosurgical board examinations. These studies compared the results achieved by different AI models with one another, as well as with the performance of medical students, residents, and neurosurgeons.

In five articles,^(12–14,16,18)^ a comparative analysis was conducted among various AI chatbots (**Table 2**).

**Table 2.** Comparative Performance of AI Chatbots in Neurosurgical Question-Answering Tasks.

| Study | Question Count and Type | % Accuracy of AI Chatbots |  |  |  |
| --- | --- | --- | --- | --- | --- |
|  |  | GPT-3.5 | GPT-4 | Bard* | Bing** |
| Ali *et al.*<sup>(12)</sup> | 500 TB questions (389 TB and 111 IB) | 73.4 | 83.4 | N/I | N/I |
|  | 111 IB questions | 49.5 | 56.8 |  |  |
| Ali *et al.*<sup>(13)</sup> | 149 TB questions (98 TB and 51 IB) | 62.4 | 82.6 | 44.2 | N/I |
|  | 51 IB questions | 47.1*** | 68.6*** | 66.7<sup>†</sup> |  |
| Guerra *et al.*<sup>(14)</sup> | 643 questions (477 TB and 166 IB) | 58.7<sup>††</sup> | 76.6<sup>†††</sup> | N/I | N/I |
|  | 477 TB questions | 60.2 | 79 |  |  |
| Stengel *et al.*<sup>(16)</sup> | 86 questions (75 TB and 11 IB) | 58 | N/I | 62 | 53 |
|  | 75 TB questions | 65 |  | 69 | 60 |
| Liu *et al.*<sup>(18)</sup> | 50 TB questions | 42 | 72 | N/I | N/I |

TB: Text-based. IB: Image-based. N/I: Not included in the study. \*Rebranded as Gemini. \*\*Rebranded as Microsoft Copilot. \*\*\*Only answered 47/51 questions. <sup>†</sup>Only answered 49/51 questions. <sup>††</sup>Only answered 618/643 questions.
<sup>†††</sup>Only answered 591/643 questions.

The comparative analysis of AI chatbots reveals significant advancements in their capabilities, particularly with newer iterations like GPT-4, which consistently outperformed earlier versions such as GPT-3.5 (also known as ChatGPT). This improvement is especially evident in handling complex theoretical questions, reflecting enhanced reasoning and contextual understanding. However, all chatbots, including GPT-4, demonstrated notable limitations in addressing image-based tasks, highlighting a persistent challenge in multimodal reasoning.

The variability in performance across chatbots, such as Bard (Gemini) and Bing (Copilot), underscores the influence of architecture and training data on their effectiveness. While Bard showed competitive results in certain contexts, Bing consistently underperformed, suggesting gaps in its integration of real-time data and reasoning capabilities. These findings emphasize the potential of AI chatbots as valuable tools for theoretical training but also reveal critical gaps in their ability to handle practical, image-based, and open-ended clinical scenarios. Addressing these limitations through enriched multimodal datasets and standardized evaluation protocols will be essential for their responsible integration into neurosurgical education.

Seven articles^(12,14–19)^ examined the performance of these chatbots in comparison to medical students, residents, and neurosurgeons (**Table 3**).

**Table 3.** Comparative Performance of AI Chatbots vs. Students, Residents, and Neurosurgeons.

| Study | Questions | Human<br>Participants | % of Accuracy |  |  |  |  |
| --- | --- | --- | --- | --- | --- | --- | --- |
|  |  |  | Human (Mean %) | GPT-3.5 | GPT-4 | Bard* | Bing** |
| Ali *et al.*<sup>(12)</sup> | 500 TB questions (389 TB and 111 IB) | Historical data from past residents (n = N/R) | 73.7 | Refer to Table 2. |  |  |  |
| Guerra *et al.*<sup>(14)</sup>,<br>Hopkins *et al.*<sup>(15)</sup> | 643 questions (477 TB and 166 IB) | Students (n = 4) | 26.3 | 58.7*** | 76.6<sup>†</sup> | N/I | N/I |
|  |  | Residents (n = 2) | 61.5 |  |  |  |  |
| Stengel *et al.*<sup>(16)</sup> | 86 questions (75 TB and 11 IB) | Historical data from past residents (n = N/R) | 59 | Refer to Table 2. |  |  |  |
|  | 75 TB questions |  |  |  |  |  |  |
| Sahin *et al.*<sup>(17)</sup> | 523 TB questions | Historical data from past residents (n = 260) | 62.02 | 78.77 | N/I | N/I | N/I |
| Liu *et al.*<sup>(18)</sup> | 50 TB questions | Low seniority neurosurgeons (n = 3) | 40 | Refer to Table 2. |  |  |  |
|  |  | Middle seniority<br>neurosurgeons<br>(n = 3) | 46 |  |  |  |  |
|  |  | High seniority<br>neurosurgeons<br>(n = 3) | 78 |  |  |  |  |
| Bartoli *et al.*<sup>(19)</sup> | 50 questions (43 TB and 7 IB) | Residents (n = 10) | 52,4 | 50,8<sup>††</sup> | N/I | N/I | N/I |

TB: Text-based. IB: Image-based. N/I: Not included in the study. N/R: Not reported in the study. \*Rebranded as Gemini. \*\*Rebranded as Microsoft Copilot. \*\*\*Only answered 618/643 questions. †Only answered 591/643 questions. ††Only answered 42/50 questions.

Advanced chatbots, such as GPT-4, consistently outperformed earlier iterations and human counterparts in theoretical questions, even surpassing junior neurosurgeons in certain domains. However, their accuracy declined markedly in image-based queries or scenarios requiring clinical interpretation, where trainees and practitioners retained significant advantages.

Performance disparities between chatbots and humans also varied by expertise level. While chatbots matched or exceeded students and residents in theoretical tasks, their results were less consistent against senior neurosurgeons, who maintained superiority in complex, integrated clinical decision-making. This gap underscores the irreplaceable role of hands-on experience and clinical intuition in neurosurgical training, areas where AI currently falls short.

GPT-4 demonstrates marked superiority over GPT-3.5 across neurosurgical subspecialties, particularly excelling in structured domains like functional and vascular neurosurgery (**Table 4**). Its enhanced performance reflects iterative improvements in contextual reasoning and specialized knowledge integration. However, variability in complex or interpretation-heavy areas, such as trauma and pediatrics, underscores lingering challenges in handling clinical ambiguity. These advancements position GPT-4 as a valuable tool for theoretical training, though its limitations in nuanced decision-making reaffirm the irreplaceable role of clinical expertise and human judgment.

**Table 4.** Comparative Performance of GPT-3.5 vs. GPT-4 on Neurosurgical Areas.

| Study | GPT-3.5 vs. GPT-4's % of Accuracy |  |  |  |  |  |  |  |
| --- | --- | --- | --- | --- | --- | --- | --- | --- |
|  | Functional | Pain | Pediatrics | PN | Spine | Trauma | Tumor | Vascular |
| Ali *et al.*<sup>(12)</sup> | 81 vs.<br>95.2 | 53.3 vs.<br>73.3 | 73.2 vs.<br>80.4 | 100 vs.<br>100 | 81.5 vs.<br>93.5 | 79.7 vs.<br>87.5 | 74.6 vs.<br>92.5 | 64 vs.<br>72 |
| Ali *et al.*<sup>(13)</sup> | 87.5 vs.<br>87.5 | N/R | N/R | 55 vs.<br>75 | 64.3 vs.<br>90.5 | N/R | 52.4 vs.<br>71.4 | 68 vs.<br>84 |
| Guerra *et al.*<sup>(14)</sup>, Hopkins *et al.*<sup>(15)</sup> | 70.5<br>vs.<br>83.39 | 69.1<br>vs.<br>84 | 68.8<br>vs.<br>78.5 | 69.1<br>vs.<br>84 | 56.3<br>vs.<br>73 | 67.6<br>vs.<br>77.8 | 59<br>vs.<br>75 | 62.1<br>vs.<br>81 |
| Liu *et al.*<sup>(18)</sup> | 37 vs.<br>77.7 | N/R | N/R | N/R | N/R | 77.7 vs.<br>89 | 71.7 vs.<br>86.7 | 49 vs.<br>89 |
| **Mean % of Accuracy** | 69 vs.<br>85.9 | 61.2 vs.<br>78.85 | 71 vs.<br>79.45 | 74.7 vs.<br>86.3 | 67.4 vs.<br>85.7 | 75 vs.<br>84.7 | 64.4 vs.<br>81.4 | 60.8 vs.<br>81.5 |

PN: Peripheral nerve. N/R: Not reported in the study.

Ethically, chatbots’ success in theoretical assessments raises concerns about overreliance in clinical decision-making or unsupervised academic preparation. For instance, their tendency to generate inaccurate responses to image-based questions could propagate errors if used without expert validation. Furthermore, excessive dependency on chatbots risks undermining trainees’ acquisition of critical diagnostic reasoning and therapeutic prioritization skills.

To mitigate these risks, chatbots should be integrated as supplementary educational tools rather than standalone solutions, focusing on reinforcing theoretical knowledge and literature review. Future research must prioritize enhancing their multimodal data-processing capabilities (e.g., radiological images) and establishing governance frameworks to regulate their ethical use in clinical and academic settings.

### Neurosurgical Simulation-Based Training and Tutoring

In fives articles,^(20–24)^ AI algorithms or techniques were employed in skills training via surgical simulation technologies (**Table 5**).

**Table 5.**
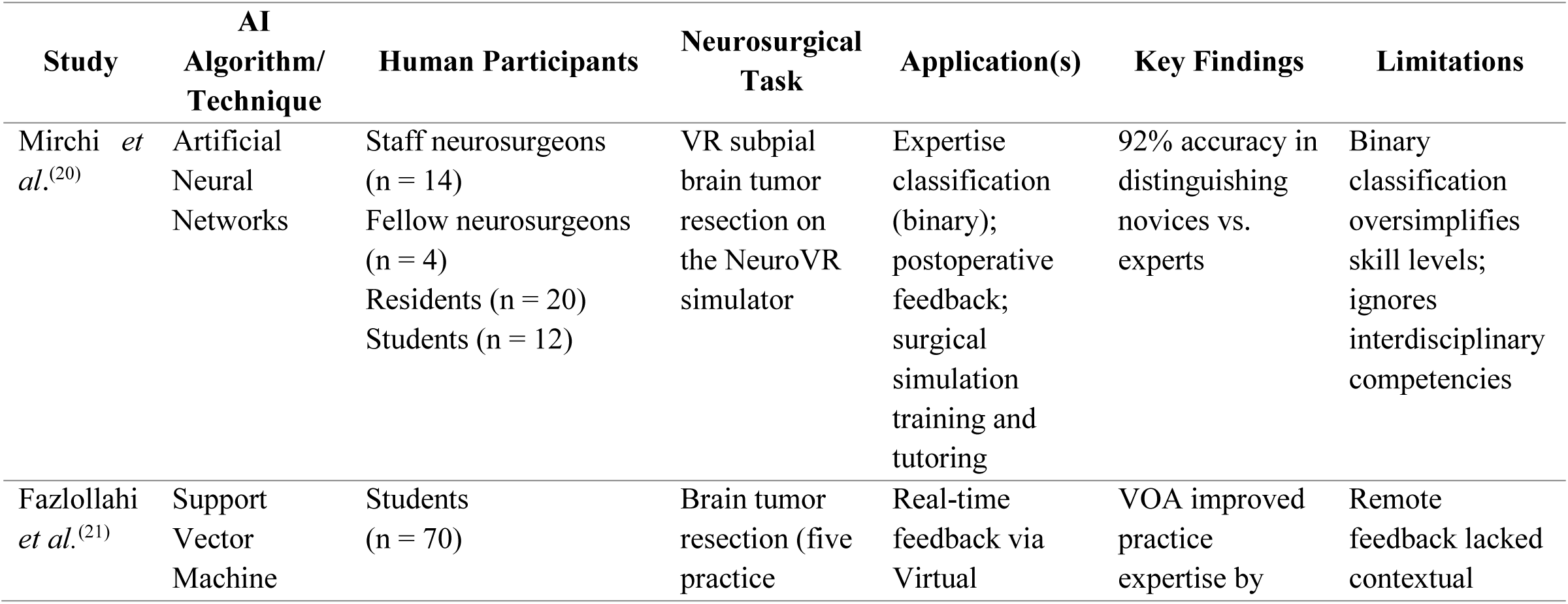
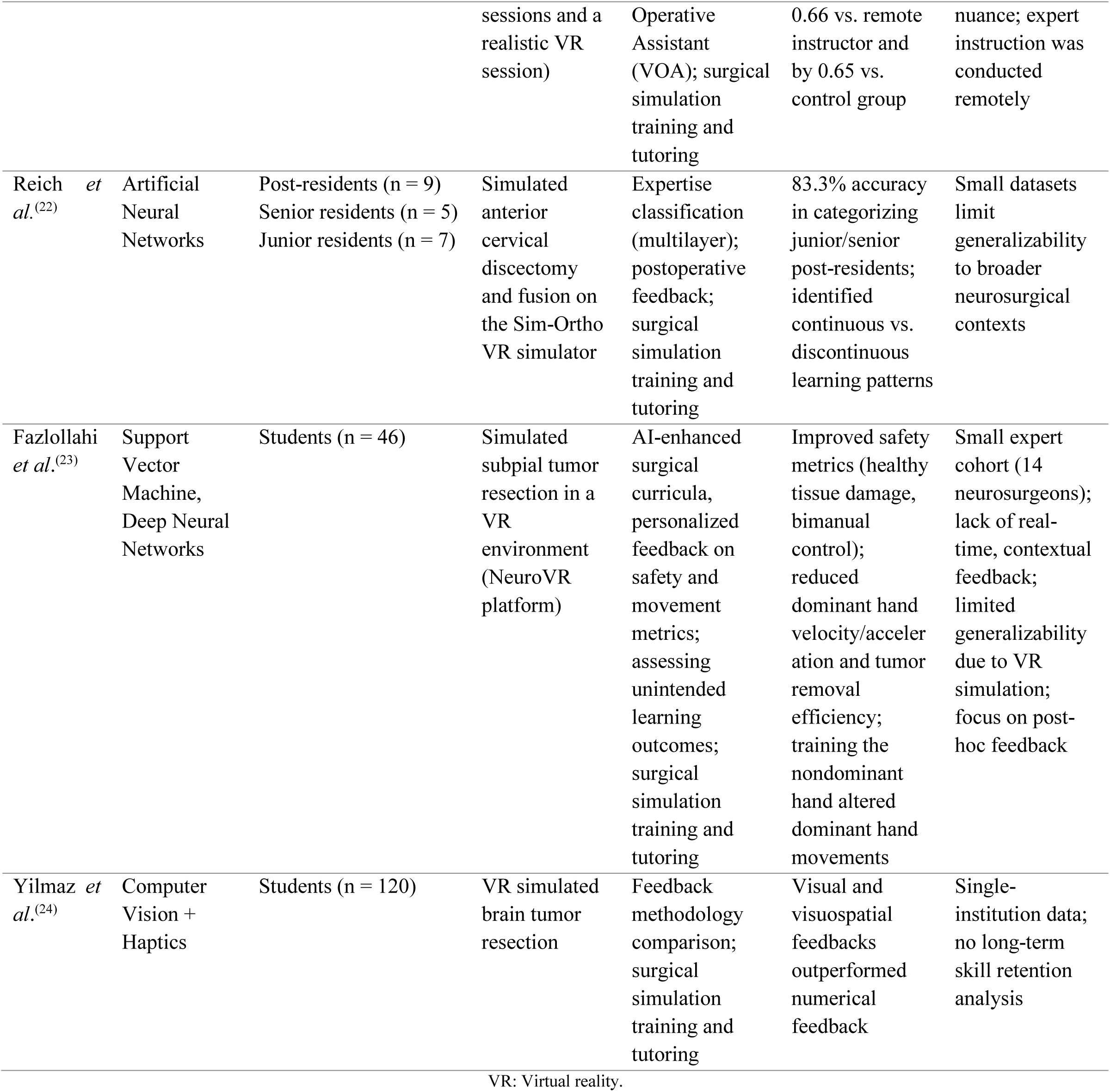
Applications in Neurosurgical Simulation-Based Training and Tutoring.

The integration of AI into neurosurgical simulation training demonstrates a dual capacity to enhance technical skill acquisition while exposing critical gaps in holistic surgical education. AI-driven systems, such as those using Support Vector Machines (SVM),^(21)^ excel in standardizing feedback focused on quantifiable metrics, such as instrument efficiency and safety parameters. However, these systems risk oversimplifying skill progression through binary classifications,^(20)^ which fail to capture nonlinear learning trajectories. For example, Reich *et al*.^(22)^ observed confidence-driven risk-taking behaviors in senior residents, contrasting with the cautious precision of post-residents, illustrating the complexity of skill development that rigid classifications may overlook.

Hybrid models merging AI-driven analytics with expert mentorship emerge as pivotal solutions. While AI outperformed remote instructors in improving objective performance scores,^(21)^ human feedback proved superior in addressing qualitative aspects like operative strategy and instrument handling. This highlights the complementary roles of quantitative precision and qualitative mentorship. However, the reliance on remote instruction during the pandemic underscored limitations in replicating the depth of in-person mentorship, particularly in delivering nuanced contextual feedback.

The modality and timing of feedback significantly shape training outcomes. Visuospatial guidance,^(24)^ for instance, proved more effective than numerical methods in refining bimanual control, though the long-term retention of these skills remains unstudied. Post-hoc AI feedback models, while effective in optimizing safety metrics, inadvertently reduced procedural efficiency in tasks like tumor removal, revealing unintended consequences of prioritizing isolated metrics over holistic performance.^(23)^

Further analysis reveals critical nuances. Mirchi *et al*.^(20)^ emphasize the risk of metric compensation bias, where strong performance in one skill may obscure deficiencies in others, necessitating transparent, multi-dimensional evaluations. Reich *et al*.^(22)^ identify nonlinear learning patterns tied to trainee experience, such as shifts in confidence and risk tolerance, which demand adaptive, experience-tailored training frameworks. Additionally, AI models consistently overlook non-technical competencies, such as interdisciplinary collaboration and ethical decision-making, which remain vital for comprehensive surgical expertise.

Future advancements should prioritize hybrid curricula that harmonize AI’s granular analytical capabilities with expert-led contextual mentorship. Expanding validation efforts to real-world, longitudinal contexts will be essential to assess skill retention and adaptability across diverse surgical scenarios. Addressing gaps in interdisciplinary skill integration and ensuring balanced curriculum design —one that avoids over-prioritizing metrics at the expense of procedural efficiency— will solidify AI’s role as a transformative yet complementary tool in neurosurgical education.

### Neurosurgical Performance/Skills/Expertise Analysis and Assessment

Seven studies^(25–31)^ investigated the role of AI algorithms/techniques in assessing the performance of students, residents, and neurosurgeons in simulated procedures (**Table 6**).

**Table 6.**
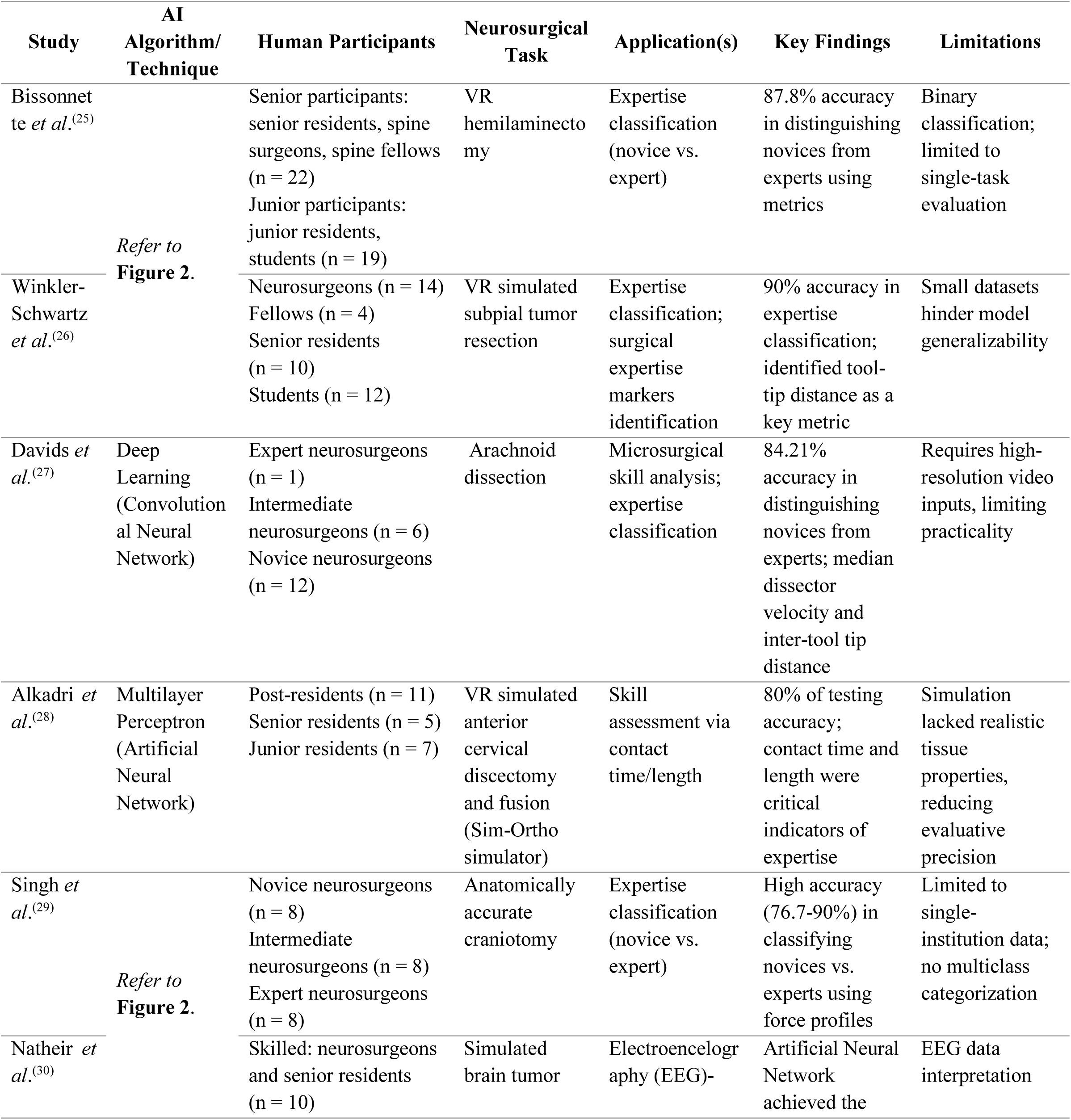
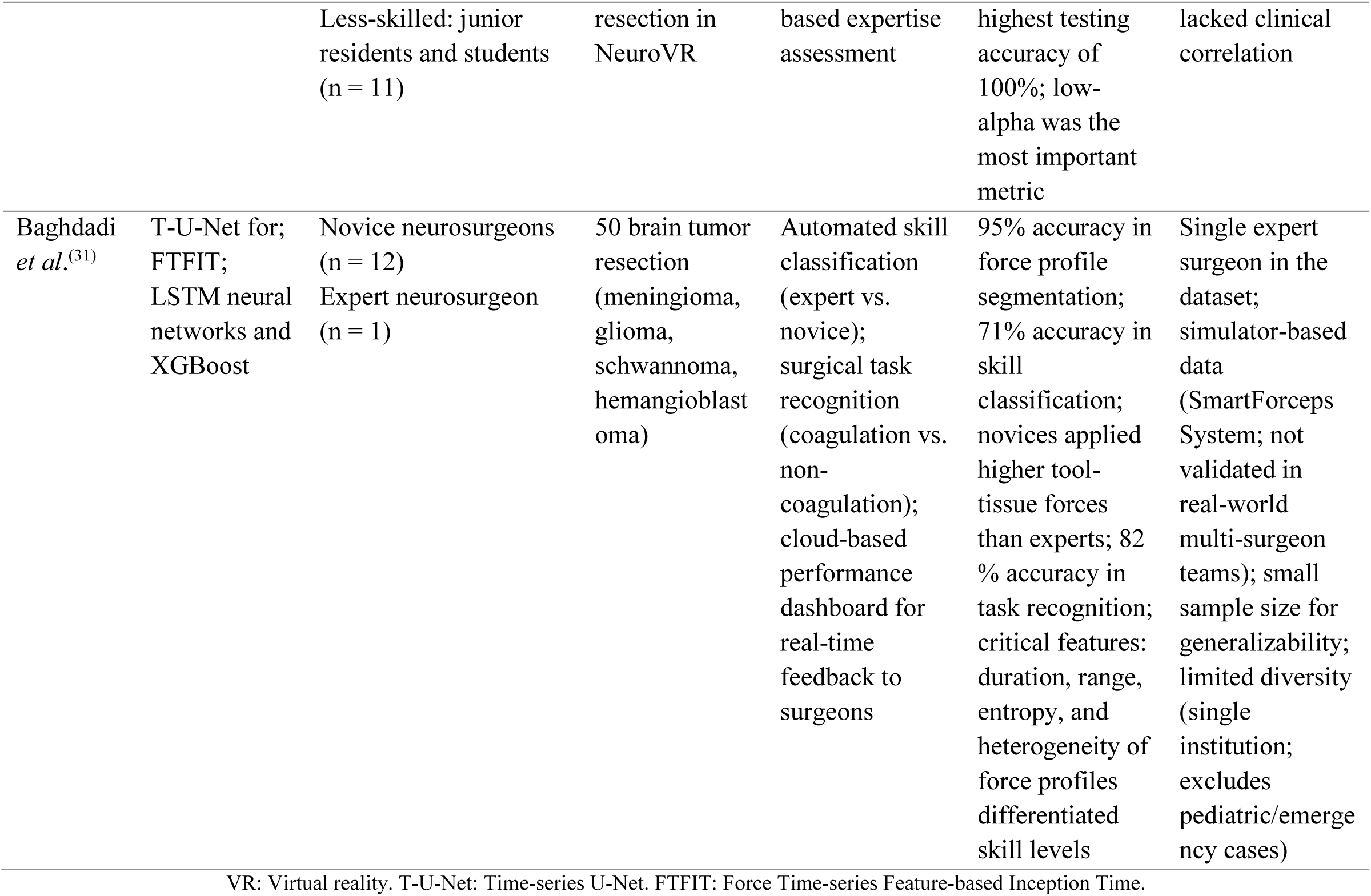
Applications in Neurosurgical Performance/Skills/Expertise Analysis and Assessment.

Four of these articles^(25,26,29,30)^ compared the accuracy of various ML algorithms in correctly assessing each participant’s expertise level: SVM, Linear Discriminant Analysis (LDA), Naïve Bayes (NB),^(25,26,29,30)^ K-Nearest Neighbors (KNN),^(25,26,30)^ Decision Tree (DT),^(25,29)^ Artificial Neural Network (ANN), Logistic Regression (LR), and Random Forest (RF)^(30)^ (**Figure 2**). Notably, the LDA model demonstrated high precision, ranking among the top three models with the highest accuracy in three of the four articles,^(25,26,29)^ with accuracy values of 87.8%, 78%, and 81.9%, respectively. Following this, the SVM,^(25,30)^ NB,^(26,29)^ and KNN^(25,26)^ models appeared twice among the top three models with the highest accuracy.

**Figure 2.**
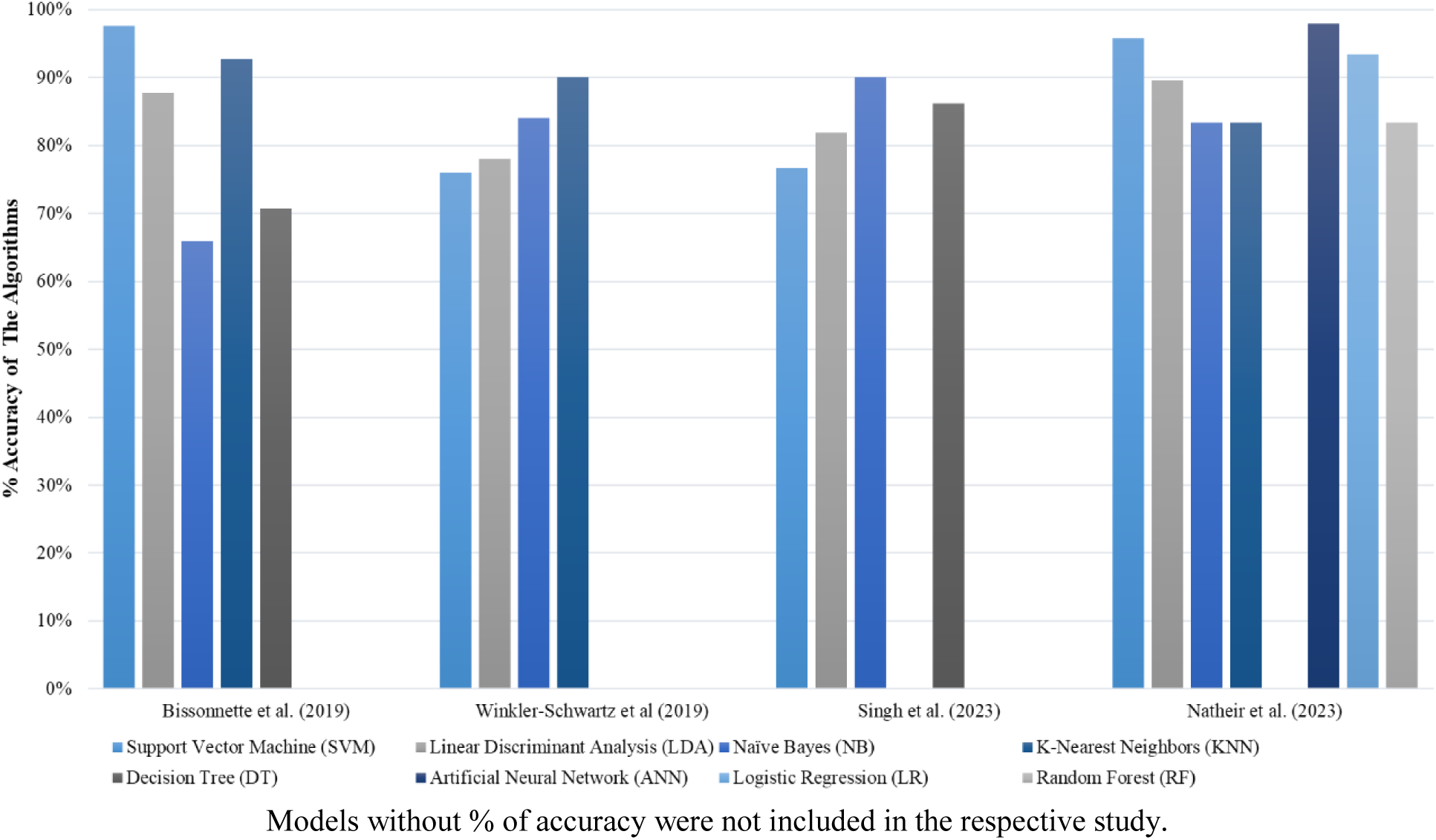
Accuracy of ML Algorithms Used for Assessing The Participant’s Expertise Level.

These findings highlight the potential of AI integration into surgical assessment systems, proposing a hybrid evaluation model where AI aids trainees in skill development tracking, performance benchmarking, and targeted feedback tailored to proficiency levels.^(28)^ However, limitations such as small, institution-specific datasets restrict model generalizability and adaptability to new contexts.^(26, 28)^ To address this, future iterations should prioritize expanded datasets encompassing diverse procedures and expertise levels. Additionally, enhancing simulation realism through biomechanically accurate tissue models^(29)^ and diverse surgical instrument integration^(25)^ would improve evaluative precision, ensuring assessments better reflect real-world surgical competency. Together, these advancements could bridge gaps between AI-driven metrics and holistic skill development, fostering robust, adaptable training frameworks.

Additionally, four studies^(25,27,28,31)^ utilized AI-provided feedback during participant evaluations to describe specific behavioral differences, linking these differences to the participants’ levels of experience. Similarly, three articles^(25,27,31)^ describe behavior trends in different participant groups at various stages of simulated surgery.

### Other Applications in Neurosurgical Education

In addition to the applications already discussed, three articles^(32–34)^ highlighted other uses of AI, showcasing its great potential in neurosurgical education.

Sevgi *et al.*^(32)^ evaluated ChatGPT’s utility in generating educational content, such as clinical cases and multiple-choice questions (MCQs). The AI produced three MCQs, two of which were correct and complete, while one contained two correct answers —a critical error in exam settings. ChatGPT also generated two clinically coherent case studies: one involving meningitis complicated by an epidural hematoma post-lumbar puncture and another detailing endoscopic lumbar discectomy for leg pain. While the format and content of the cases were deemed appropriate, the study emphasized the need for expert oversight to refine outputs and ensure quality. The authors suggest that with improved model training and domain-specific databases, AI could become a valuable tool for developing neurosurgical question banks and standardized exams for trainees.^(32)^

Witten *et al.*^(33)^ developed a convolutional neural network (CNN)-based algorithm to segment neuroanatomical structures from 879 images sourced from *The Neurosurgical Atlas*. The model, trained on manually labeled categories (e.g., arteries, veins, brain tissue), employed data augmentation (rotation, cropping, blurring) to enhance generalization. It achieved 91.8% test accuracy, with intersection-over-union (IoU) scores varying by tissue type: 0.887 for arteries and 0.674 for brain tissue. Lower performance on brain tissue was attributed to imprecise manual labeling. The authors propose integrating this segmentation model with augmented reality or virtual simulation technologies to provide real-time anatomical feedback during training, improving residents’ ability to identify and manipulate structures preoperatively.^(33)^

Balu *et al.*^(34)^ explored surgical instrument detection using the YOLOv4 model on video frames from endonasal simulations of internal carotid artery injury repairs (SOCAL database). The model achieved 59.4% accuracy overall, rising to 74.3% for metallic instruments due to higher contrast. By calculating Shannon entropy (ShEn) —a measure of instrument variability— the study linked procedural success to instrument use patterns. Successful procedures exhibited higher entropy (ShEn = 0.452 vs. 0.370 for unsuccessful cases). A logistic regression model predicted success with 73.9–78.3% accuracy using automated detections. The findings suggest that analyzing instrument patterns could help design structured educational frameworks to teach efficient surgical workflows, though entropy metrics may vary in complex, heterogeneous procedures.^(34)^

These studies collectively underscore AI’s expanding role in neurosurgical education, from generating training materials to enhancing anatomical visualization and procedural analytics. However, challenges such as data quality, model generalizability, and the need for human-AI collaboration remain critical to ensuring reliable and scalable implementations. Future advancements in AI, coupled with expert validation, could transform how neurosurgeons acquire and refine technical and cognitive skills.

### Limitations

This scoping review has limitations. The broad inclusion criteria led to significant heterogeneity in study designs and outcomes, making it difficult to draw definitive conclusions. The narrative synthesis approach, while providing a rich contextual understanding, limited quantitative aggregation and comparability of results. Additionally, the review may have missed relevant studies due to the constraints of the database, search strategy, language restrictions, selection criteria, and the unavailability of full texts. The absence of a formal risk of bias assessment means the findings should be interpreted with caution. Finally, given the rapid advancements in AI technology, the review’s findings may quickly become outdated, necessitating continuous updates to stay current with the evolving field.

## CONCLUSIONS

The results of this scoping review highlight the significant potential of AI in neurosurgical education. AI models have demonstrated significant benefits in enhancing board examination performance, providing valuable feedback during simulation-based training, accurately assessing surgical skills, and offering innovative educational tools. However, the findings underscore the importance of addressing ethical considerations, utilizing comprehensive datasets, and ensuring realistic simulation scenarios.

Moving forward, the continuous development and refinement of AI models, coupled with collaboration between educators, AI developers, and policymakers, will be essential to maximize the effective and responsible integration of AI in neurosurgical training.

## Data Availability

All data produced in the present study are available upon reasonable request to the authors.

## LIST OF ABBREVIATIONS

AI: Artificial Intelligence
PRISMA: Preferred Reporting Items for Systematic Reviews and Meta-Analyses
PRISMA-ScR: Preferred Reporting Items for Systematic Reviews and Meta-Analyses extension for Scoping Reviews
PRISMA-A: Preferred Reporting Items for Systematic Reviews and Meta-Analyses extension for Abstracts
SVM: Support Vector Machines
LDA: Linear Discriminant Analysis
NB: Naïve Bayes
KNN: K-Nearest Neighbors
DT: Decision Tree
ANN: Artificial Neural Network
LR: Logistic Regression
RF: Random Forest
CNN: Convolutional Neural Network
IoU: Intersection-over-Union
YOLOv4: You Only Look Once version 4
EEG: Electroencephalography
VR: Virtual Reality

## DECLARATIONS

### Ethics Approval and Consent to Participate

Not applicable.

### Consent for Publication

Not applicable.

### Availability of Data and Materials

No datasets were generated or analyzed during the current study.

### Author’s Contributions

Conceptualization: Hector Julio Piñera-Castro.

Data curation: Hector Julio Piñera-Castro, Christian Borges-García.

Formal analysis: Hector Julio Piñera-Castro, Christian Borges-García.

Investigation: Hector Julio Piñera-Castro, Christian Borges-García.

Methodology: Hector Julio Piñera-Castro.

Resources: Hector Julio Piñera-Castro.

Supervision: Hector Julio Piñera-Castro.

Visualization: Hector Julio Piñera-Castro, Christian Borges-García.

Writing – original draft: Hector Julio Piñera-Castro, Christian Borges-García.

Writing – review & editing: Hector Julio Piñera-Castro, Christian Borges-García.

